# Modeling the epidemiological impact of different adult pneumococcal vaccination strategies in the United Kingdom

**DOI:** 10.1101/2024.10.21.24315757

**Authors:** Rachel J Oidtman, Giulio Meleleo, Oluwaseun Sharomi, Ian R Matthews, Dionysios Ntais, Robert Nachbar, Tufail M Malik, Kevin M Bakker

## Abstract

**Background:** Pneumococcal conjugate vaccines (PCVs) were first introduced in the pediatric United Kingdom (UK) immunisation programme in 2006 which led to significant declines in invasive pneumococcal disease (IPD) caused by targeted serotypes. Although pediatric PCVs provide some indirect protection to adults, a significant IPD burden remains in older adults. Here, we compared three adult (65+ years-old) and risk group (2-64-year-old) vaccination scenarios, namely a continuation of the status quo with PPSV23 vaccination, using the recently licensed-in-adults PCV20, or using the new adult-focused 21-valent PCV, V116.

**Methods:** A population-level compartmental dynamic transmission model (DTM) was adapted to the UK setting. The model described *Streptococcus pneumoniae* carriage transmission dynamics and disease progression in the presence of age- and serotype-specific pneumococcal vaccines. We calibrated the DTM to age- and serotype-specific IPD data in the UK and used the model to make projections under the different adult vaccination scenarios, while keeping PCV13 immunization in children.

**Results:** The calibrated model yielded reasonable parameter values and model fits that closely matched observed IPD dynamics. Among 65+ year-olds, routine use of V116 averted more cases of IPD than PCV20 or PPSV23 vaccination. There was a notable decrease in IPD incidence in the serotypes unique to V116. In the serotypes included in PCV20 but not V116, the model did not predict a resurgence of IPD.

**Conclusions:** Projections revealed that in adults, V116 led to greater reductions in IPD in the 65+ year-old population compared with PCV20 or PPSV23.

**HIGHLIGHTS:**

- A dynamic transmission model was able to replicate historical pneumococcal dynamics.
- An adult specific PCV (V116) in the UK would avert more pneumococcal disease than other candidate adult vaccines.
- The dynamic model predicted no resurgence of serotypes not included in V116.

## INTRODUCTION

*Streptococcus pneumoniae* is an important pathogen and leading cause of vaccine-preventable bacterial infections. Infants and the elderly population are the most vulnerable. Invasive pneumococcal diseases (IPD), such as meningitis and bacteremic pneumonia, are rare but can lead to high levels of morbidity and mortality, while non-invasive pneumococcal diseases are more common but cause less mortality [1, 2]. There are more than 100 immunologically distinct *S. pneumoniae* serotypes with different serotypes exhibiting different life history traits, such as invasiveness (i.e., ability to cause pneumococcal disease) across age groups [3, 4].

In 2003 the United Kingdom (UK) included a pneumococcal polysaccharide vaccine (PPSV23) as a recommendation to the adult vaccination schedule, while two pneumococcal conjugate vaccines (PCVs), PCV7 then PCV13, were recommended for children in 2006 and 2010, respectively [5]. Although the routine use of PCVs in pediatrics has led to concomitant declines in vaccine-type serotypes (VTs) in pediatrics due to direct protection and adults due to indirect protection [6], a significant burden of adult IPD remains in serotypes not included in PCV13, including 8, 10A, 11A, 12F, 15A, 15C, 16F, 23A, 23B, 24F, 31, and 35B [7]. This enduring burden underscores the need for a PCV designed specifically for adults.

A new adult-focused 21-valent PCV, V116, has been developed to protect against serotypes that predominantly circulate in adults, including eight serotypes that have not previously been included in a pneumococcal vaccine (15A, 15C, 16F, 23A, 23B, 24F, 31, 35B) [8]. Meanwhile, a 20-valent PCV (PCV20) developed for pediatrics was recently approved for use in adults in the UK. PCV20 includes 15 serotypes covered by previous PCVs and five additional serotypes (8, 10A, 11A, 12F, 15B) [9]. In 65+ year-olds in the UK in 2019, 88% of IPD was caused by serotypes included in V116 while 61% of IPD was caused by serotypes included in PCV20 [10].

The rich history of pneumococcal vaccination in the UK has caused multiple indirect effects, including herd protection and serotype replacement. Therefore, dynamic transmission models (DTMs) – mathematical models of infectious disease dynamics that can reproduce direct and indirect effects – are recommended [11] to answer our questions regarding the impact of new pneumococcal vaccines [12]. An important feature of *S. pneumoniae* ecology is that following the introduction of a new PCV, VTs decrease in prevalence and are sometimes replaced by non-vaccine types (NVTs) [10, 13]. This replacement is driven by competition between resident and invading serotypes and has previously been incorporated into DTMs of *S. pneumoniae* transmission in the UK [14–18] and other settings [19–32].

Here, we investigated the impact of different adult pneumococcal vaccination scenarios on the incidence of pneumococcal disease outcomes in the UK. A previously described DTM [33] was adapted to the UK setting and calibrated to historical age- and serotype-specific IPD data in the presence of historical vaccinations. We used the calibrated model to project IPD under continued use of PPSV23 compared to V116 vaccination versus PCV20 vaccination.

## METHODS

### Data

The Joint Committee on Vaccination and Immunisation (JCVI) in the UK recommended PPSV23 in 1992 for hisk-risk individuals 2 years of age and older and extended this to a broader age-based recommendation in 2003 for individuals 80 years of age and older [5]. By 2005, PPSV23 eligibility had been extended to all individuals aged 65 years of age and older and all individuals in clinical risk groups (such as individuals with diabetes, immunosuppression, or chronic heart disease) [5]. In the pediatric population, PCV7 was introduced into the National Immunisation Program in a 2+1 schedule for the 0-1- year-old population in 2006 and was later replaced by PCV13 in 2010 [5]. A 1+1 pediatric vaccination schedule began in 2020 [5] but given the lack of real-world data for updated vaccine effectiveness under a reduced dosing schedule, we did not explicitly consider a 1+1 dosing schedule in model projections. Data on vaccination coverage rates by age were available from the UK Health Security Agency (UKHSA) [34] (Table S4-S6). Given the high vaccination coverage rate (VCR) in PPSV23 starting in 2005, we assumed that from 2000-2005 the PPSV23 coverage increased linearly from 0% to the target VCR of 64% (Table S5). Vaccination coverage rates by risk group were available from the UKHSA PPSV Coverage Report [35], with which we calculated the weighted VCR across all risk groups combined to estimate a constant population-level VCR for 2-64-year-olds (Table S6).

Annual age- and serotype-specific IPD incidence data for 2000-2019 were provided by the UKHSA [10]. To align the model projected IPD incidence to observed IPD incidence from 2000-2019, we first assumed that the steady state represented the historical *S. pneumoniae* dynamics in the absence of widespread pneumococcal vaccines. Therefore, the model’s steady state was calibrated to the average of the 2000- 2005 IPD incidence data by age and serotype. For the pre-PCV era, we assumed that serotype-specific *S. pneumoniae* carriage data from 2006 [13] was a representative baseline target for the model. Specifically, we considered overall carriage ranges and *S. pneumoniae* carriage serotype distribution for 0-5-year-olds from Cleary et al. [13] as a model calibration target (Figure S1) and assumed that the pediatric *S. pneumoniae* carriage serotype distribution applied to 5-64- and 65+ year-olds. For 5-64- and 65+ year-olds we used Hussain et al. [36] as a model calibration target for overall *S. pneumoniae* carriage (Figure S2). For the model fitting, we aggregated serotype-specific IPD and carriage data into 11 serotype classes (STCs) (Table 1).

**Table 1.** Serotype classes (STCs) with respective serotypes (STs) included in each class. Shading indicates inclusion in different vaccines. V116 contains deOAc15B pneumococcal polysaccharide which has a molecular structure similar to 15C (referred to as 15C hereafter). Note that serotype 3 was included in a stand-alone STC due to evidence of limited immunogenicity response from vaccines [56].

| Serotype class (STC) | Serotypes included in class | Vaccination Group | PCV7 | PCV13 | PCV15 | PCV20 | V116 | PPSV |
| --- | --- | --- | --- | --- | --- | --- | --- | --- |
| 1 | 4, 6B, 9V, 14, 18C, 19F, 23F | PCV7 | - | - | - | - |  | - |
| 2 | 1, 5 | PCV13-non-PCV7-non-ST3 |  | - | - | - |  | - |
| 3 | 3 | ST3 |  | - | - | - | - | - |
| 4 | 7F, 19A | PCV13-non-PCV7-non-ST3 |  | - | - | - | - | - |
| 5 | 6A, 6C | PCV13-non-PCV7-non-ST3 |  | - | - | - | - |  |
| 6 | 22F, 33F | PCV15-non-PCV13 |  |  | - | - | - | - |
| 7 | 9N, 17F, 20 | PPSV23-non-PCV20 |  |  |  |  | - | - |
| 8 | 8, 10A, 11A, 12F | PCV20-non-PCV15 |  |  |  | - | - | - |
| 9 | 15B | PCV20-non-PCV15 |  |  |  | - | - | - |
| 10 | 15A, 15C, 16F, 23A, 23B, 24F, 31, 35B | Unique V116 STs |  |  |  |  | - |  |
| 11 | Non-vaccine types (NVTs) | NVTs |  |  |  |  |  |  |

Data from clinical trials, real-world evidence, and other published sources were used as inputs into our model. Demographic data, including age-specific population data, births, and deaths were from the UK Office for National Statistics [37, 38]. Data on age-specific contact rates were from a well-established mixing matrix [39]. Serotype and age-based data on vaccine effectiveness against disease were available for PCVs [40] and PPSV23 [41]. Given the lack of real-world evidence for vaccine effectiveness against disease for novel vaccine serotypes, we assumed equivalency with previous VTs in adult PCVs (Table S8). Data on serotype-specific carriage clearance rates were available from studies from multiple countries and settings [42–54], which were averaged over serotype and age to obtain aggregate inputs into the model (Table S3).

### Model description and calibration

We adapted a published dynamic transmission model [33] to the UK by updating demographic and epidemiological inputs (Table S1-S3). Broadly the model describes the *Streptococcus pneumoniae* carriage transmission dynamics among individuals and disease progression in the presence of age- and serotype-specific pneumococcal vaccines.

Given the lack of detailed data to parameterize the model for all possible serotypes and to reduce model complexity, we grouped serotypes into 11 STCs based on their inclusion in different vaccines (Table 1) [55]. We aggregated serotype-specific data into STCs and assumed that serotypes within the same STC and age group were characterized by the same model parameter values. Broadly, the model includes two core components: a demographic model and a transmission model.

#### Demographic model

The demographic model encompassed population size and age structures. In brief, to best account for patterns of pneumococcal carriage, differences in disease manifestations, vaccination recommendations in the UK, and available data in the UK, the total population was divided into four age groups: 0-1-, 2-4-, 5-64-, and 65+ years-old. Within the demographic model, there was aging, represented by a maturation rate into older age groups (Table S1). There were also births and deaths within the model, including a fertility rate and a probability of death from natural causes within each age group which are formulated such that there was a closed population (Table S1).

Given the different age groups within the population, we accounted for heterogeneity in contact rates due to different mixing patterns among age groups. Mixing among different age groups is particularly important with *S. pneumoniae* as infants and young children act as a pathogen reservoir [44, 57]. We transformed a well-established mixing matrix for 5-year age increments [39] to obtain a mixing matrix for the four modeled age groups in the UK (Table S2). The mixing matrix was then implemented in the transmission model.

#### Transmission model

The transmission model distinguished between carriage and disease and explicitly modeled the transmission of *S. pneumoniae* carriage via a set of differential equations [33]. The progression from carriage to pneumococcal disease was modeled through an age-, serotype-, and vaccine-status-specific relationship, wherein disease occurs in some fraction of carriage (Figure 1).

**Figure 1.**
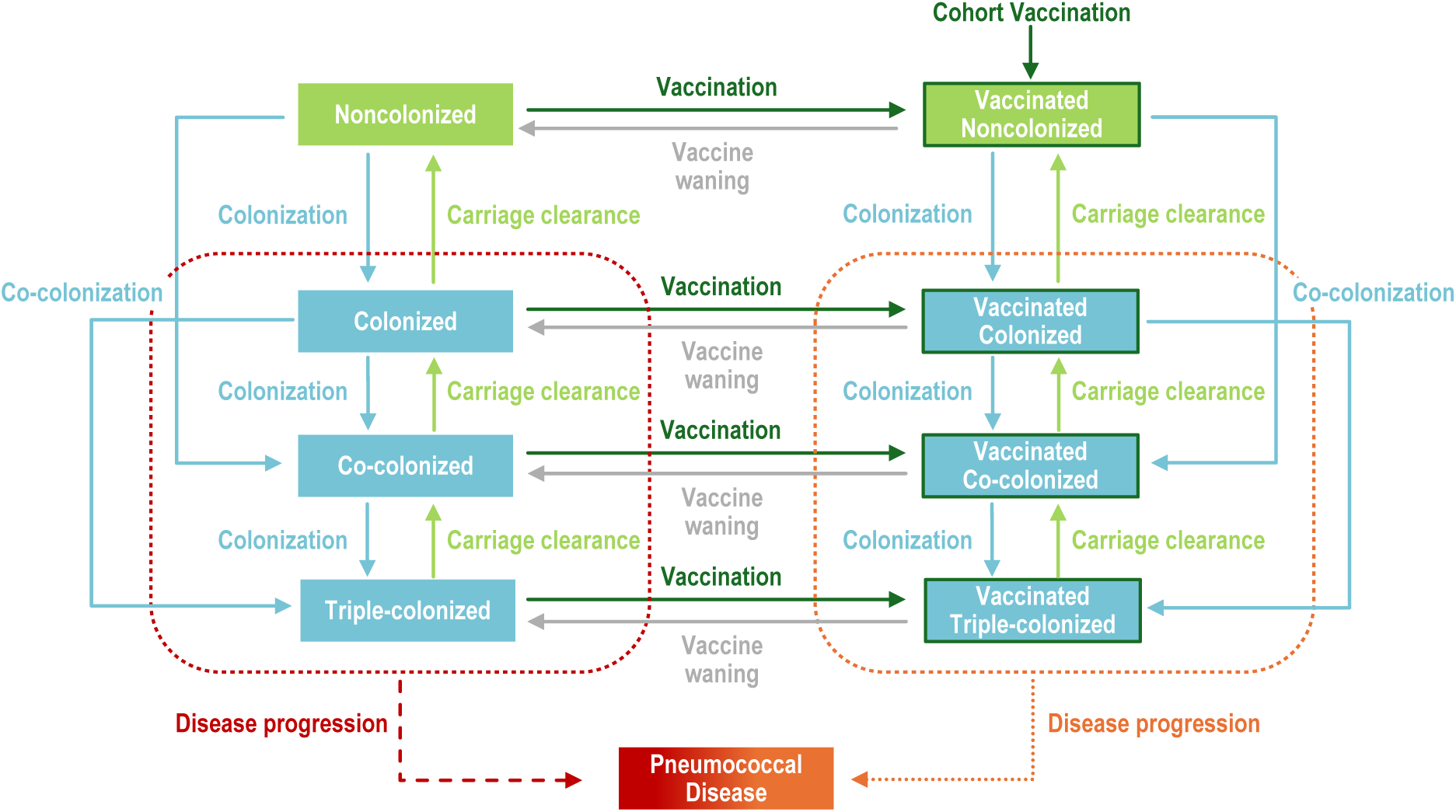
Model diagram. Upon carriage acquisition, individuals moved to either the colonized, co-colonized, or triple-colonized classes, depending on whether one or two serotypes were acquired (denoted in blue). Some portion of individuals from these colonized classes would develop a pneumococcal disease (denoted in red and orange). Recovery then moved individuals back to the non-colonized class (denoted in light green), sequentially clearing one serotype at a time. Vaccination (dark green border) reduced the risk of carriage acquisition and disease development for vaccine serotypes.

Within the epidemiological model, there were non-colonized individuals, singly colonized individuals, co-colonized (i.e., colonized by two STCs simultaneously), and triple-colonized (i.e., colonized by three STCs simultaneously) individuals. Within each of these classes, individuals could be unvaccinated or vaccinated with a PCV vaccine, a PPSV vaccine, or a PCV and PPSV vaccine. The rate at which individuals became colonized was STC- and age-specific (i.e., transmission rate) and was reduced by the vaccine efficacy against carriage for the respective STCs. The vaccine efficacy waning (i.e., its durability) varied dependent on vaccine. Competition from one colonizing STC could also reduce the risk of secondary colonization by an invading STC (i.e., competition). The STC- and age-specific clearance rate upon colonization (i.e., recovery rate) was the reciprocal of the average duration of carriage and did not vary based on vaccination status. Upon colonization with STC(s) an individual could develop pneumococcal disease (e.g., IPD) at STC-, age-, and disease-specific rates, which was reduced in vaccinated individuals due to the vaccine efficacy against disease.

We used data to inform serotype-specific vaccine efficacy against IPD (i.e., vaccine efficacy against disease) and estimated vaccine efficacy against carriage. In the calibration period, vaccine efficacies against disease for PCV7, PCV13, and PPSV23 were included (Table S7-S8). For projections, we assumed continued PCV13 vaccination in pediatrics. In adults and risk groups, due to the lack of real-world evidence, new VT serotypes in V116 and PCV20 were assumed to have similar levels of protection as previous serotypes in adult PCVs (Table S8). For shared serotypes between PCV20 and V116, the VEs are identical within age groups.

#### Calibration

First, we calibrated the model’s projected VCR to the vaccination coverage data to ensure that it captured the different vaccination introductions and increases in vaccination coverage over time (Figure S6). Time-varying cohort and continuous vaccination rates were calibrated so the vaccination coverage values in the model matched historical data across the different age groups (Table S4-S6). For model projections we assumed VCRs in pediatrics, adults, and risk groups continued at the 2019 VCR levels. For PCVs, for all age groups, we assumed a mean duration of protection of 10 years based on Leidner et al. 2021 [58]. For PPSV23, for all relevant age groups, we assumed a mean duration of protection of 7.5 years based on Bonten et al. 2015 [59].

We calibrated the transmission model to age- and STC-specific IPD incidence per 100,000 (referred to as IPD incidence from hereon) data from the UK. Estimated parameters include vaccine efficacy against carriage, carriage acquisition rate given contact, competition between STCs, and invasiveness (case-to-carrier ratio) (Table S10-S13). Each of these calibrated values were age- and STC-specific except for competition, which was STC-STC specific. For the competition parameters, we used historical replacement dynamics to calibrate the specific STC-STC combinations. In the absence of observed replacement dynamics, the competition parameters were calibrated to 1 (i.e., no explicit competition). The model was calibrated using the Nelder-Mead simplex method to minimize a weighted sum of squared error’s objective function over the annual age- and STC-specific IPD incidence data. The Nelder-Mead simplex method implemented in the NMinimize function in Mathematica 13.3.1 was used for calibration [60].

#### Vaccination scenarios

The calibrated model was used to evaluate the impact of three vaccines (PPSV23, V116, and PCV20) on the adult and risk group population, while maintaining consistency in the pediatric vaccine across all vaccine scenarios.

The three scenarios are summarized below:

1. **Base case scenario with PPSV23 vaccination**: Adults aged 65+ years-old and clinical risk groups aged 2-64-years-old received PPSV23 at assumed vaccination coverage rates. Children aged 0-1-years-old receive PCV13 at assumed vaccination coverage rates.
2. **Vaccination with V116**: Adults aged 65+ years-old and clinical risk groups aged 2-64-years-old received V116 at assumed vaccination coverage rates. Children aged 0-1-years-old receive PCV13 at assumed vaccination coverage rates.
3. **Vaccination with PCV20**: Adults aged 65+ years-old and clinical risk groups aged 2-64-years-old received PCV20 at assumed vaccination coverage rates. Children aged 0-1-years-old receive PCV13 at assumed vaccination coverage rates.

#### Sensitivity analysis

To assess the degree to which uncertainty in specific model parameters affected the epidemiological outcomes (IPD incidence) in our projections, we performed a one-way deterministic sensitivity analysis (DSA). In the DSA, we varied the duration of protection in both adult PCVs and the vaccine efficacy in both adult PCVs (Table S9). We considered three broad scenarios: base case VE against disease, low VE against disease, and high VE against disease, each with a base case duration of protection, short duration of protection, or long duration of protection, for a total of nine scenarios for both V116 and PCV20.

## RESULTS

### Model calibration

For each of the STC and age-group combinations, we assessed the model’s fit to the data. The model was able to replicate the decline in incidence in new VT STCs following the introduction of vaccines, such as the dramatic declines in incidence of PCV13 serotypes following the introduction of pediatric PCV13 vaccination in 2010 (Figure 2, middle column). Additionally, the model reproduced the replacement dynamics in PCV15-non-PCV13 and unique V116 serotypes (Figure 2, middle and right column) following the declines of PCV13 serotypes. The model was not able to fully replicate some of the rapid increases in PCV20-non-PCV15 and PPSV23-non-PCV20 serotypes in 5-64- and 65+ year-olds starting in 2015 (Figure 2, middle and right column). Overall, the calibrated parameters (Table S10-S13) yielded IPD incidence levels that closely followed the observed IPD incidence levels (Figure S3) during the calibration period. Post hoc, we calculated the root mean square error (RMSE) to evaluate the STC- and age-specific predictions of annual incidence to observed STC- and age-specific annual incidence [61] (Figure S4), wherein an RMSE closer to zero indicates a better fit while a higher value indicates more error. By age, the RMSE values were 1.47, 0.597, 0.312, and 1.29 for the 0-1, 2-4, 5-64, and 65+ age groups. Across all age groups, the RMSE was 1.03.

**Figure 2.**
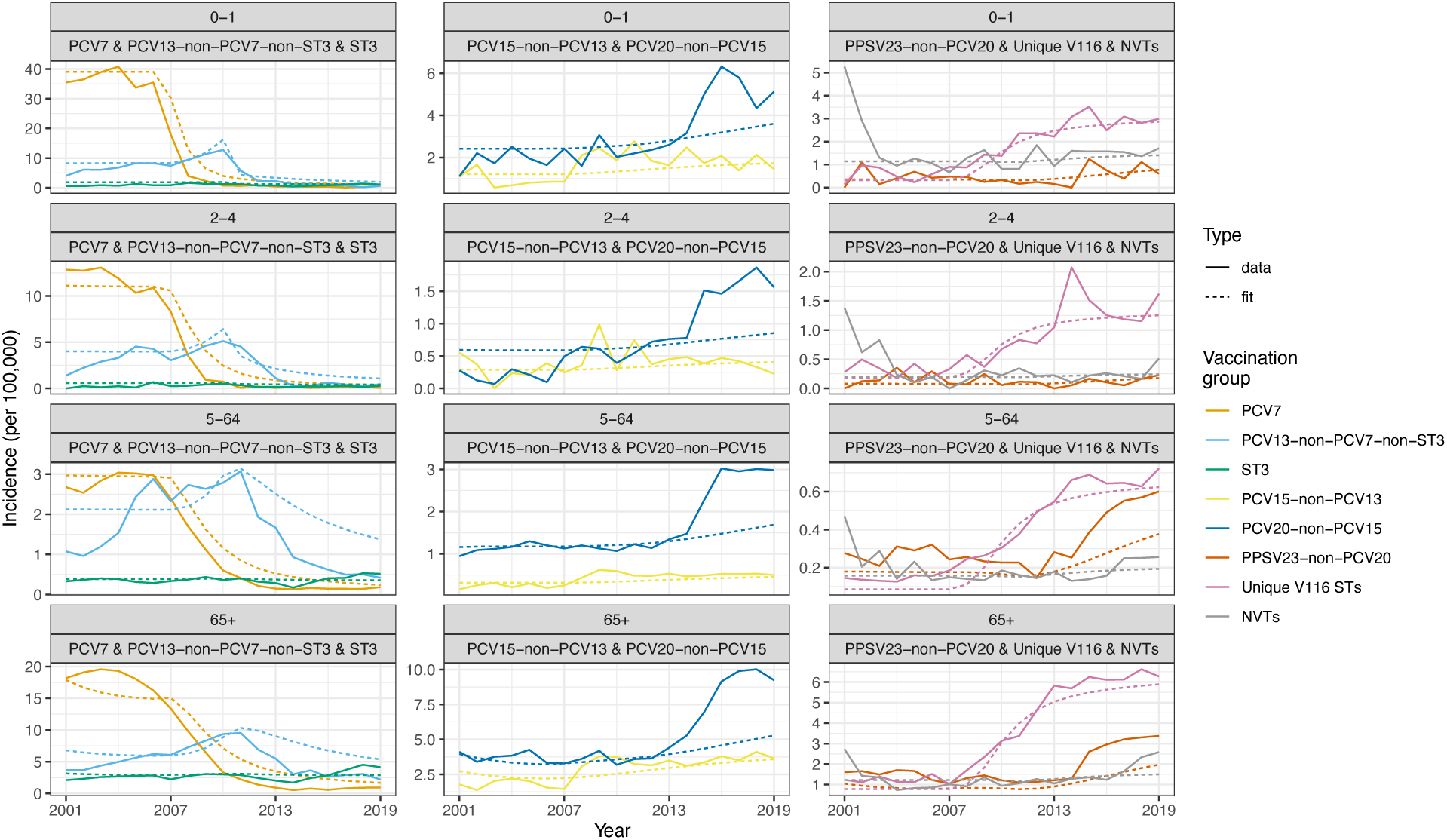
Model calibration compared to data. The correspondence between empirical IPD incidence (solid line) and modeled IPD incidence (dotted line) by vaccination group and age group.

### Projections under different vaccination scenarios

We assessed the epidemiological impact of V116 or PCV20 vaccination in at risk 2-64-year-olds and 65+ year-olds compared to the base case of PPSV23 vaccination in the UK. Over a 10-year vaccination time horizon, IPD incidence in the 65+ year-old population increased by 3.83% with continued PPSV23 vaccination and was reduced by 15.5% and 8.87% with V116 and PCV20 vaccination, respectively (Table 2, Figure S5). Over the same 10-year projection period, changes in IPD incidence due to risk group vaccination in the 2-4- and 5-64-year-old varied depending on vaccine and age group (Table 2).

**Table 2.** Projected IPD incidence. IPD incidence at the start of the projection and 10 years after the introduction of adult (in 65+ year-olds) and risk group vaccination (in 2-64-year-olds) with PPSV23, V116, or PCV20. Incidence is stratified by vaccinated age group.

| Scenario | Age group | IPD incidence at start of projection | 10-year projections of IPD incidence (% change from projection start) |
| --- | --- | --- | --- |
| Base case (PPSV23) | 2-4 | 4.81 | 4.65 (-3.40%) |
| V116 | 2-4 |  | 4.51 (-6.28%) |
| PCV20 | 2-4 |  | 4.52 (-6.09%) |
| Base case (PPSV23) | 5-64 | 5.31 | 5.52 (3.83%) |
| V116 | 5-64 |  | 5.14 (-3.23%) |
| PCV20 | 5-64 |  | 5.17 (-2.67%) |
| Base case (PPSV23) | 65+ | 28.2 | 29.2 (3.83%) |
| V116 | 65+ |  | 23.8 (-15.5%) |
| PCV20 | 65+ |  | 25.7 (-8.87%) |
| Base case (PPSV23) | Overall | 10.6 | 11.0 (3.74%) |
| V116 | Overall |  | 9.50 (-10.3%) |
| PCV20 | Overall |  | 9.94 (-6.21%) |

The projected trends in IPD incidence varied by vaccination group. In 65+ year-olds and in those serotypes that were included in V116 but not PCV20 (9N, 15A, 15C, 16F, 17F, 20A, 23A, 23B, 24F, 31, and 35B), 10-year projections predicted a 12.7% decrease in incidence with V116 vaccination but a 14.0% and 18.2% increase of incidence with PPSV23 or PCV20 vaccination, respectively (Figure 3, V116-non-PCV20). In 65+ year-olds and in those serotypes that were included in PCV20 but not V116 (1, 4, 5, 6B, 9V, 14, 18C, 19F, and 23F), 10-year projections predicted decreases in incidence under all three vaccination strategies, with projected 39.9%, 28.0%, or 60.1% decreases with PPSV23, V116, or PCV20 vaccination, respectively (Figure 3, PCV20-non-V116). In the serotypes that are common between PCV20 and V116 but not included in PPSV23, 10-year projections with either adult PCVs are similar and predict reduced disease compared to PPSV23 (Figure 3, Shared serotypes). Projections did not vary across vaccine for the NVTs (Figure 3, NVT).

**Figure 3.**
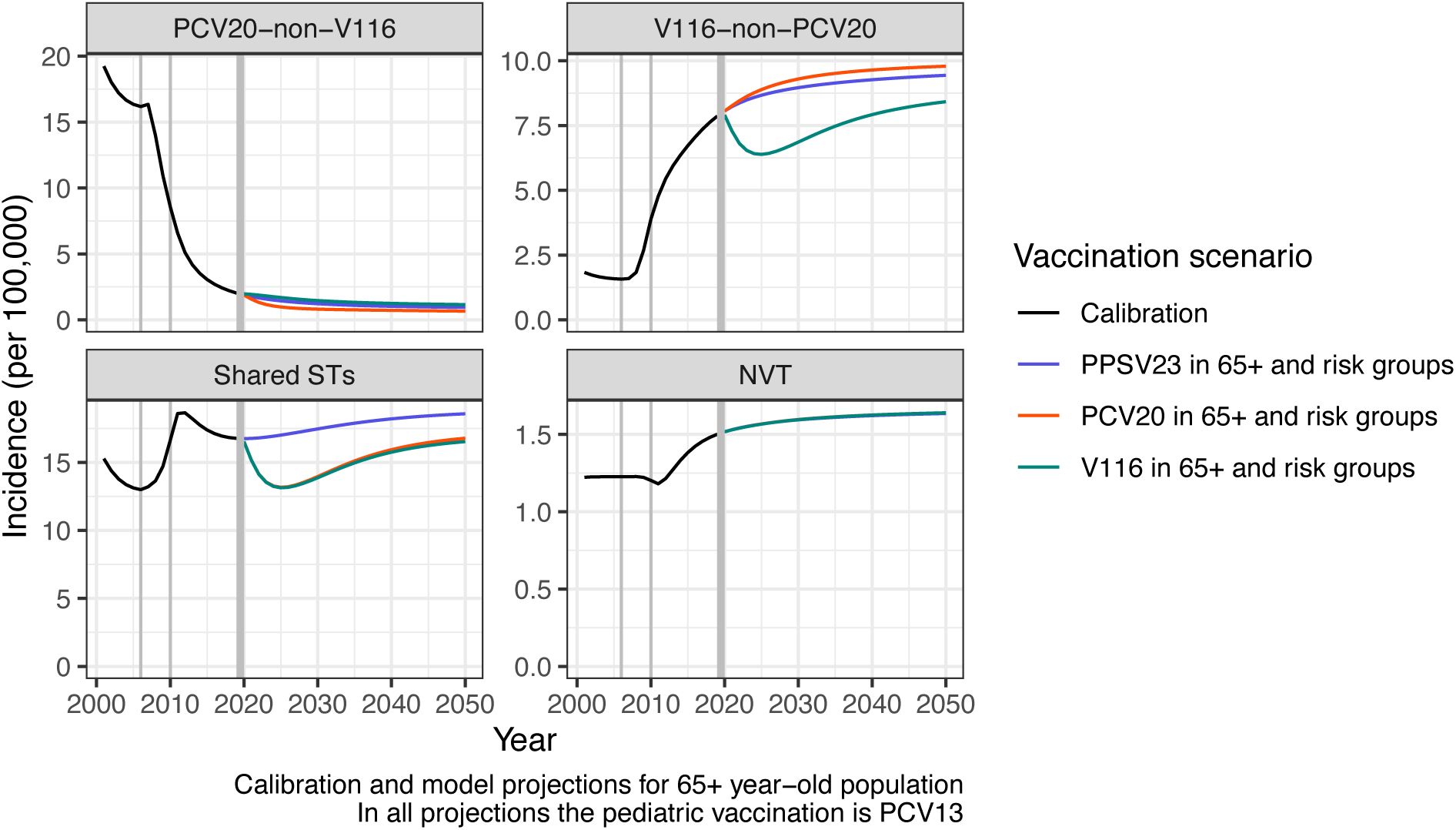
30-year projections of IPD incidence for 65+ year-olds. The three different adult vaccination scenarios for 65+ year-olds were grouped by whether the serotypes were included in PCV20 but not V116, V116 but not PCV20, both PCV20 and V116, or were non-vaccine types. The black line denotes the calibrated model. The blue line denotes a scenario with PPSV23 in 65+ year-olds and risk groups, the red line denotes a scenario with PCV20 in 65+ year-olds and risk groups, and the teal line denotes a scenario with V116 in 65+ year-olds and risk groups. All projections assumed continued PCV13 vaccination in the pediatric population. Vertical grey lines indicate when different vaccines were introduced, PCV7 in 2006, PCV13 in 2010, and new vaccinations introduced in 2020.

Risk group vaccination of 2-4-year-olds led to an associated reduction in vaccine type carriage in that age group which slightly affected carriage (and therefore disease) dynamics in 0-1-year-olds. In the scenarios with V116 or PCV20 vaccination in the 2-4-year-old risk groups, the model projected declines in IPD incidence in 0-1-year-olds, while with continued use of PPSV23 in risk groups there was no projected decline of IPD incidence in 0-1-year-olds as PPSV23 is not effective against carriage (Figure S7). Because of equivalent vaccine efficacies against carriage and disease between shared serotypes in V116 and PCV20, the ways in which changes in carriage and disease in 2-4-year-olds resulted in changes in carriage and disease in other age groups was most often consistent in shared serotypes (Figure S8). In STC 4 (which included 7F and 19A), there were slightly larger reductions in 10-year projections of IPD incidence associated with V116 vaccination (a 45.4% reduction) compared to PCV20 vaccination (a 43.0% reduction) (Figure S8).

### Sensitivity analysis

The vaccine efficacies against disease and duration of protection of newly introduced adult PCVs were varied in the DSA (Table 2). In both vaccination scenarios, the proportionally largest changes from the base case were observed with the low range parameter assumption (i.e., with low vaccine efficacy against disease and a short duration of protection) (Figure S9). Relative to the base parameter assumptions (i.e., standard duration of protection and standard vaccine efficacy against disease) projections with the low range parameter assumptions were 4.5% higher with V116 vaccination and 3.0% higher with PCV20 vaccination over the entire population (Figure S10). Overall, projection results were more sensitive to changes in vaccine efficacy against disease than changes in the duration of protection (Figure S9-S10).

The effects of varying the vaccine efficacy against disease and duration of protection for V116 and PCV20 were most apparent in the 65+ year-old population (Figure S11). Considering the extreme boundaries (i.e., low vaccine efficacy against disease + short duration of protection versus high vaccine efficacy against disease + long duration of protection), model projections still showed no overlap between the different vaccination scenarios (Figure S11), with V116 averting more disease than PCV20 or PPSV23.

## DISCUSSION

Despite more than 20 years of pneumococcal vaccination in the UK, IPD remains an important public health issue and the optimal way in which new vaccines may be used to reduce IPD needs to be determined. We calibrated an age- and serotype-specific dynamic transmission model to historical UK IPD incidence data to evaluate the epidemiological impact of different adult vaccines. The model was able to fit historical IPD dynamics, which included multiple vaccine introductions, indirect protection from pediatric vaccination, and serotype replacement caused by the introduction of new vaccines. Model projections predicted that the introduction of an adult-focused 21-valent vaccine, V116, in the 65+ year-old population and risk groups would avert more IPD cases than the continuation of PPSV23 vaccination or the introduction of PCV20 in the same population.

V116 vaccination is predicted to reduce IPD incidence to a greater extent than PCV20 and the effect was most clearly demonstrated in the serotypes that were included in V116 but not PCV20. There are 11 serotypes in V116 not included in PCV20, eight of which are serotypes that have not previously been included in a vaccine before, and therefore the inclusion in a vaccine led to concomitant declines in incidence. These serotypes have a proportionally higher burden in adults compared to pediatrics and therefore the largest reductions in IPD incidence were in the 65+ year-old population. In the 2-4-year-olds and 5-64-year-olds, the projected changes in incidence were similar with either V116 or PCV20 vaccination and led to greater reductions compared to PPSV23. As our model allowed for changes in carriage levels in 2-4-year-olds to affect carriage in 0-1-year-olds there was some variation in IPD incidence in 0-1-year-olds dependent on the risk group vaccination. Slight differences in projected incidence in shared serotypes (as was the case with STC 4) was likely related to differences in replacement dynamics and competition between serotypes. Specifically, replacement of STC 4 upon introduction of PCV20-only serotypes was greater than replacement of STC 4 upon introduction of V116- only serotypes.

As V116 is an adult-focused vaccine, it targets serotypes that primarily circulate in adult populations and therefore excludes some serotypes that historically circulate more in pediatric populations, including the original PCV7 serotypes and serotypes 1 and 5. Projections revealed a continued decline of IPD incidence in the nine serotypes excluded from V116 that were included in PCV20. This lack of resurgence in the adult population was likely due to the indirect protection provided by a robust pediatric vaccine program [62, 63], as *S. pneumoniae* transmission is driven by children and indirect protection provided by children to adults may be sufficient to stifle transmission [64, 65].

We estimated values for several parameters, including the case-to-carrier ratio and vaccine efficacy against carriage. Model estimates of the case-to-carrier ratio for PCV7 serotypes by age group closely aligned with estimates from another DTM fitted to the UK setting [14]. However these estimates vary in literature, and model estimates of the case-to-carrier ratio were one to two orders of magnitude lower than those published in Lochen et al. 2022 [66], possibly due to differences in geographic region and the types of data used to estimate the ratio. While published estimates for vaccine efficacy against carriage vary substantially [67], our estimates for pediatric vaccine efficacy against carriage for the PCV7 serotypes were within the estimated range from multiple studies [68, 69]. Further, our estimates of vaccine efficacy against carriage for serotype 3 were very low, agreeing with observations of poor immunogenicity associated with PCV13 against serotype 3 [70].

Although our model is well-calibrated and allows for the investigation of age- and serotype-specific vaccination campaigns, there are many assumptions and simplifications in the model. First, to allow for the direct estimation of vaccination impacts, the model assumed a constant population size and age structure throughout the calibration and in the projection time period, which may not be in line with projected changes in the UK population [71]. Second, like many countries around the world during the COVID-19 pandemic, the UK enacted several non-pharmaceutical interventions (NPIs) aimed at reducing the transmission of SARS-CoV-2 [72]. In the UK, these NPIs had additional downstream effects on other notifiable diseases, such as IPD [73] and other respiratory pathogens [74]. Our DTM was calibrated through 2019, and so did not include data from 2020 onward for which there is currently no mechanism in the model to include effects from the pandemic on transmission dynamics. This will be an important avenue for future model enhancements. Third, we did not incorporate the updated 1+1 pediatric dosing schedule in our projections. In future modeling work we will investigate the effects of reduced VEs associated with the reduced schedule, which will further require several assumptions to address the new dosing regimen in the absence of real-world evidence [75].

Although pediatric vaccination can reduce some adult pneumococcal disease, there remains a high burden of IPD in adults and offers an opportunity for adult-focused vaccination to alleviate this morbidity. Introducing V116, which was specifically designed for serotypes that predominantly circulate in adults, led to substantially larger predicted declines in IPD incidence in older adults than the PPSV23 and PCV20 comparators. While the results presented here are specific to the UK, they may be applicable to other similar countries considering routine adult pneumococcal vaccination.

## Data Availability

Aggregated data produced in the study are available upon reasonable request to the authors or are available in the manuscript.

## Funding

This study was funded by Merck Sharp & Dohme LLC, a subsidiary of Merck & Co., Inc., Rahway, NJ, USA.

## Declaration of Competing Interest

RO, OS, TM, and KB are full-time employees of Merck Sharp & Dohme LLC, a subsidiary of Merck & Co., Inc., Rahway, NJ, USA and may hold stock or stock options in Merck & Co., Inc., Rahway, NJ, USA. IM and DN are full-time employees of MSD (UK) Ltd, London, United Kingdom and may hold stock or stock options in Merck & Co., Inc., Rahway, NJ, USA. GM and RN are consultants whose institution (Wolfram Research, Inc., Champaign IL, USA) is paid by is paid by Merck Sharp & Dohme LLC, a subsidiary of Merck & Co., Inc., Rahway, NJ, USA. RN may hold stock or stock options in Merck & Co., Inc., Rahway, NJ, USA.

V116 was developed by Merck & Co., Inc., Rahway, NJ, USA.

## SUPPORTING INFORMATION FOR…

### Supplementary text

#### Model overview

The dynamic transmission model here was adapted from the model presented in Malik et al. [1]. The model equations are identical, including forces of carriage acquisition of one or two STCs in vaccinated or unvaccinated individuals by age and STC. Key differences include epidemiological and demographic inputs, such as the age groups: 0-1, 2-4, 5-64, and 65+, which affected the mortality hazards (Table S1), aging rates (Table S1), and mixing matrix (Table S2). Other biological constants varied because of different age groupings in the UK, specifically the duration of carriage by age group and STC (Table S3).

**Table S1.** Mortality hazards and aging rates for model age groups (28).

| Age group | 0-1 | 2-4 | 5-64 | 65+ |
| --- | --- | --- | --- | --- |
| Mortality hazards ($\mu_a$) | 0.000209 | 0.00013 | 0.0019 | 0.0506 |
| Aging rates ($m_a$) | 0.499 | 0.333 | 0.0157 | 0 |

**Table S2.** Mixing matrix for model age groups.

| Age group | 0-1 | 2-4 | 5-64 | 65+ |
| --- | --- | --- | --- | --- |
| 0-1 | 0.655 | 1.057 | 5.122 | 0.088 |
| 2-4 | 0.655 | 1.057 | 5.122 | 0.088 |
| 5-64 | 0.133 | 0.214 | 10.26 | 0.191 |
| 65+ | 0.018 | 0.029 | 3.647 | 1.128 |

**Table S3.** The estimated duration of carriage (in days) by age group and serotype class.

| Serotype Class | Serotypes in each group | 0-1 | 2-4 | 5-64 | 65+ |
| --- | --- | --- | --- | --- | --- |
| 1 | 4, 6B, 9V, 14, 18C, 19F, 23F | 70.24 | 30.60 | 24.94 | 26.83 |
| 2 | 1, 5 | 13.38 | 9.58 | 7.04 | 6.56 |
| 3 | 3 | 24.6 | 21.5 | 39.72 | 45.5 |
| 4 | 7F, 19A | 57.98 | 24.51 | 21.14 | 21.45 |
| 5 | 6A, 6C | 53.6 | 36.2 | 24.58 | 25.84 |
| 6 | 22F, 33F | 87.56 | 61.33 | 46.26 | 43.10 |
| 7 | 9N, 17F, 20 | 34.73 | 25.07 | 18.03 | 16.58 |
| 8 | 8, 10A, 11A, 12F | 39.43 | 26.59 | 20.86 | 21.47 |
| 9 | 15B | 42.24 | 31.88 | 21.32 | 21.75 |
| 10 | 15A, 15C, 16F, 23A, 23B, 24F, 31, 35B | 36.62 | 26.04 | 17.22 | 17.48 |
| 11 | NVTs | 35.26 | 30.70 | 29.32 | 31.21 |

#### Risk group vaccination

For the risk group vaccination with PCVs (either V116 or PCV20) in 2-64-year-olds, we weighted the VEs in the 2-4- and 5-64-year-old age groups to account for the fact that a significant portion of the vaccinated population (especially in the 2-4-year-old population) would still have full protection from their pediatric vaccination. We considered on one side the proportion of the vaccinated population in 2-4-year-olds that had been vaccinated with PCV13 when they were in 0-1-year-old age group and on the other side the proportion of the vaccinated population in 2-4-year-old age group that received the new PCV (either V116 or PCV20) because they were in a risk group. The weights are given by the two respective populations divided by the total of vaccinated in 2-4-year-old age group. The same argument was used for risk group vaccination in 5-64-year-old age group, since children are likely protected until 10 years old.

For the DSA we applied the same weighting to the duration of protection, considering that we only varied protection only for adult and risk group vaccination and not for pediatric vaccination. Therefore, we fixed the mean duration of protection for the population proportions in 5-64-year-olds coming from 0-1-year-old vaccination (PCV13) and we varied only the mean duration for the risk group vaccinated proportions.

### Supplementary figures

**Figure S1.**
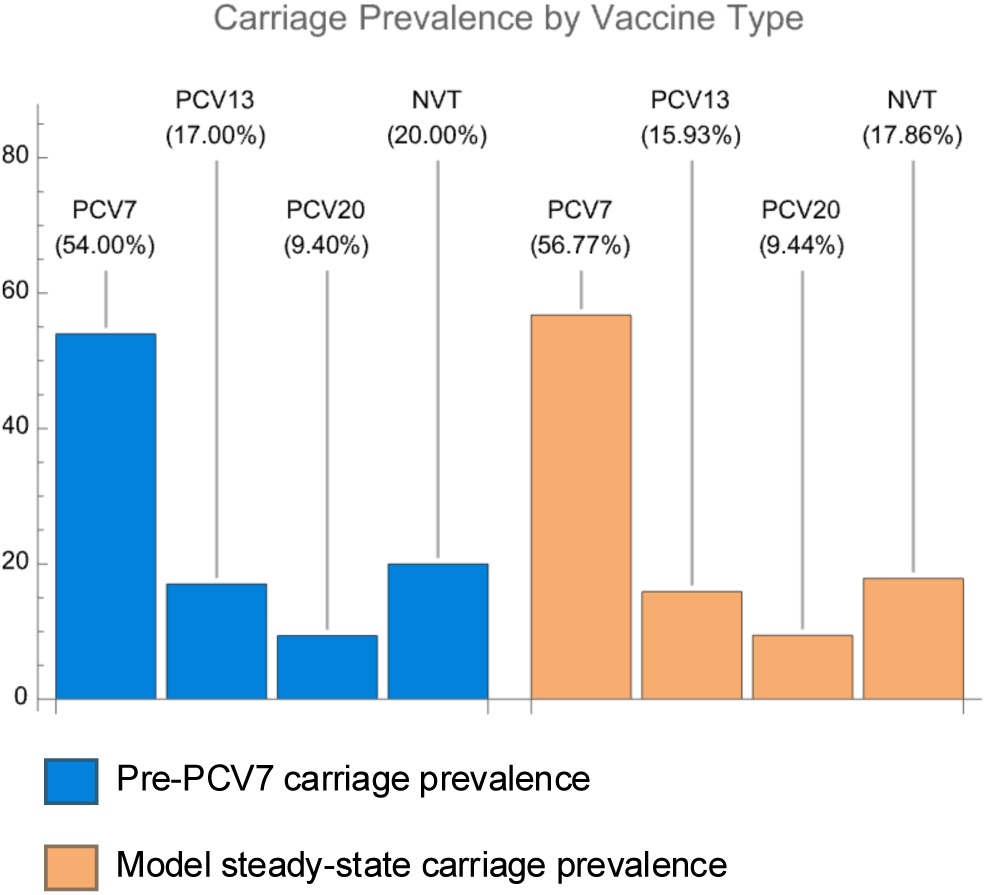
Carriage prevalence at pre-PCV steady state. Carriage prevalence by vaccination grouping from Cleary et al. [2] (blue) and from the model (orange).

**Figure S2.**
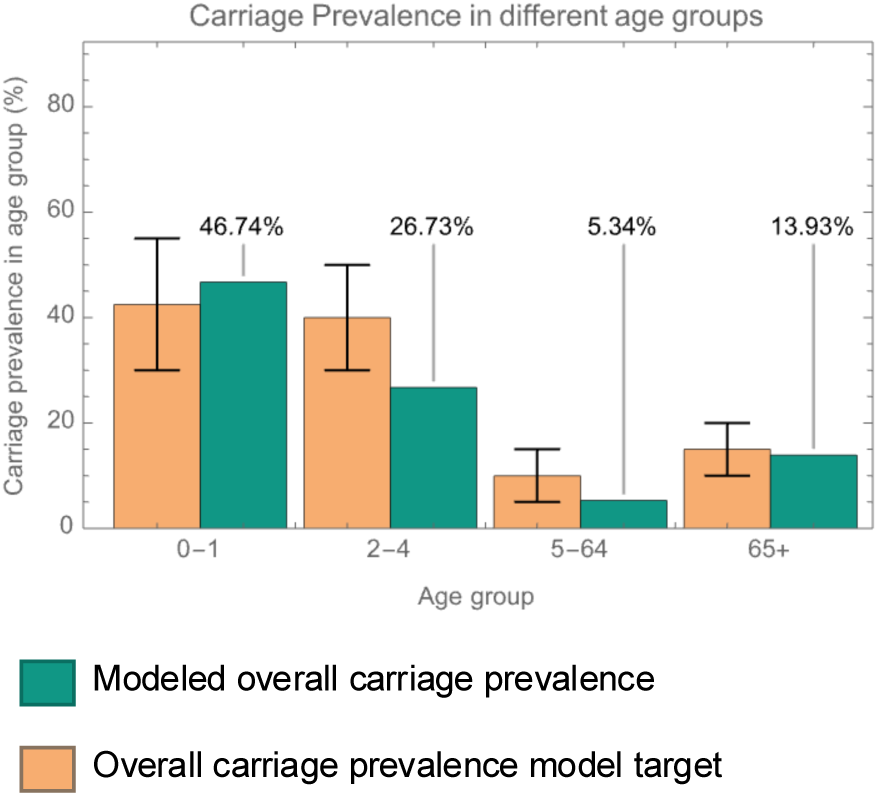
Carriage prevalence at pre-PCV steady state by age group. Carriage prevalence model calibration targets in orange were informed by Cleary et al. [2] and Hussain et al. [3]. Modeled carriage prevalence is in green.

**Figure S3.**
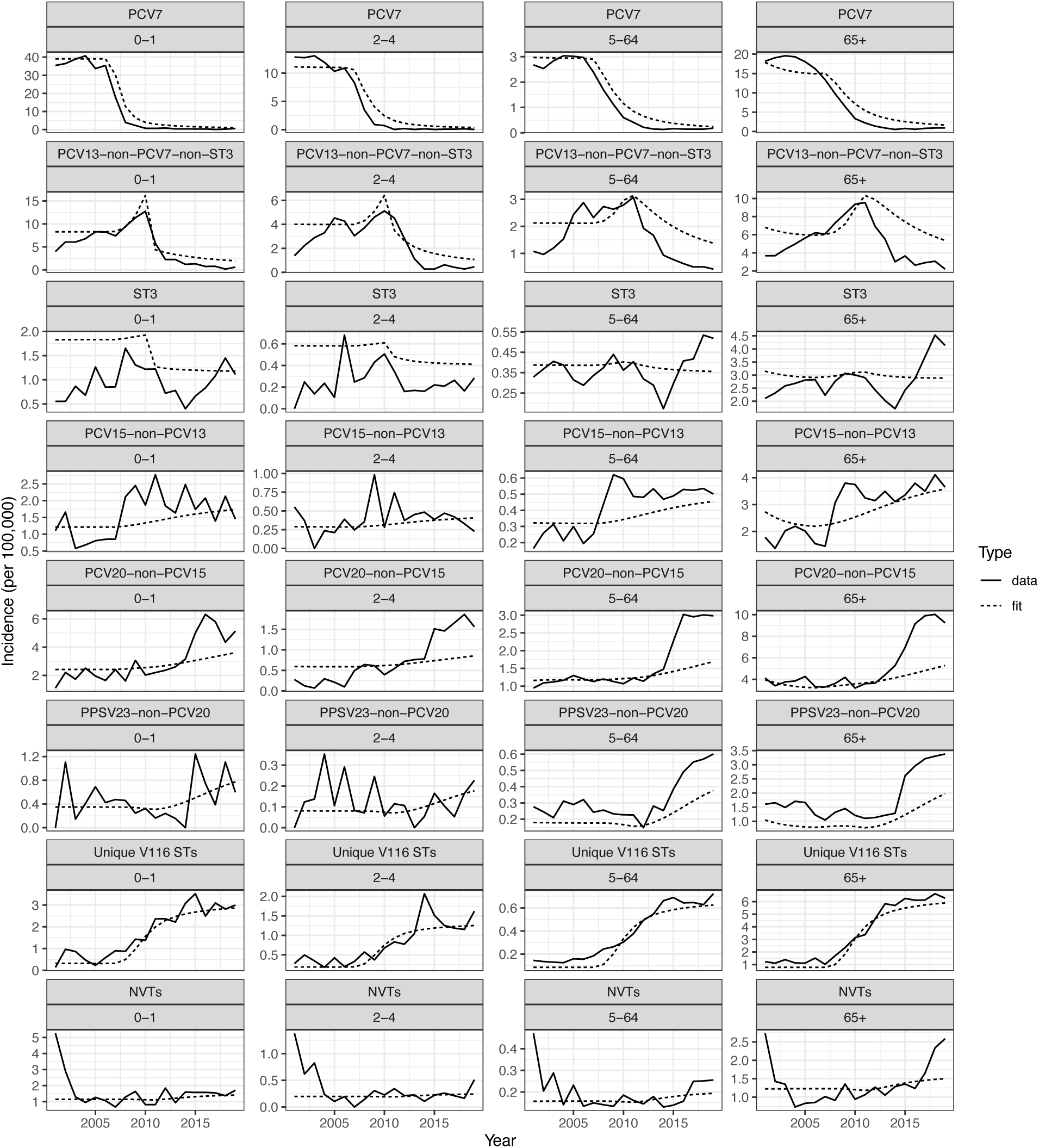
Model validation of calibrated model. We compared the data (solid line) to model fit (dotted line) during the calibration period faceted by age group and vaccination grouping.

**Figure S4.**
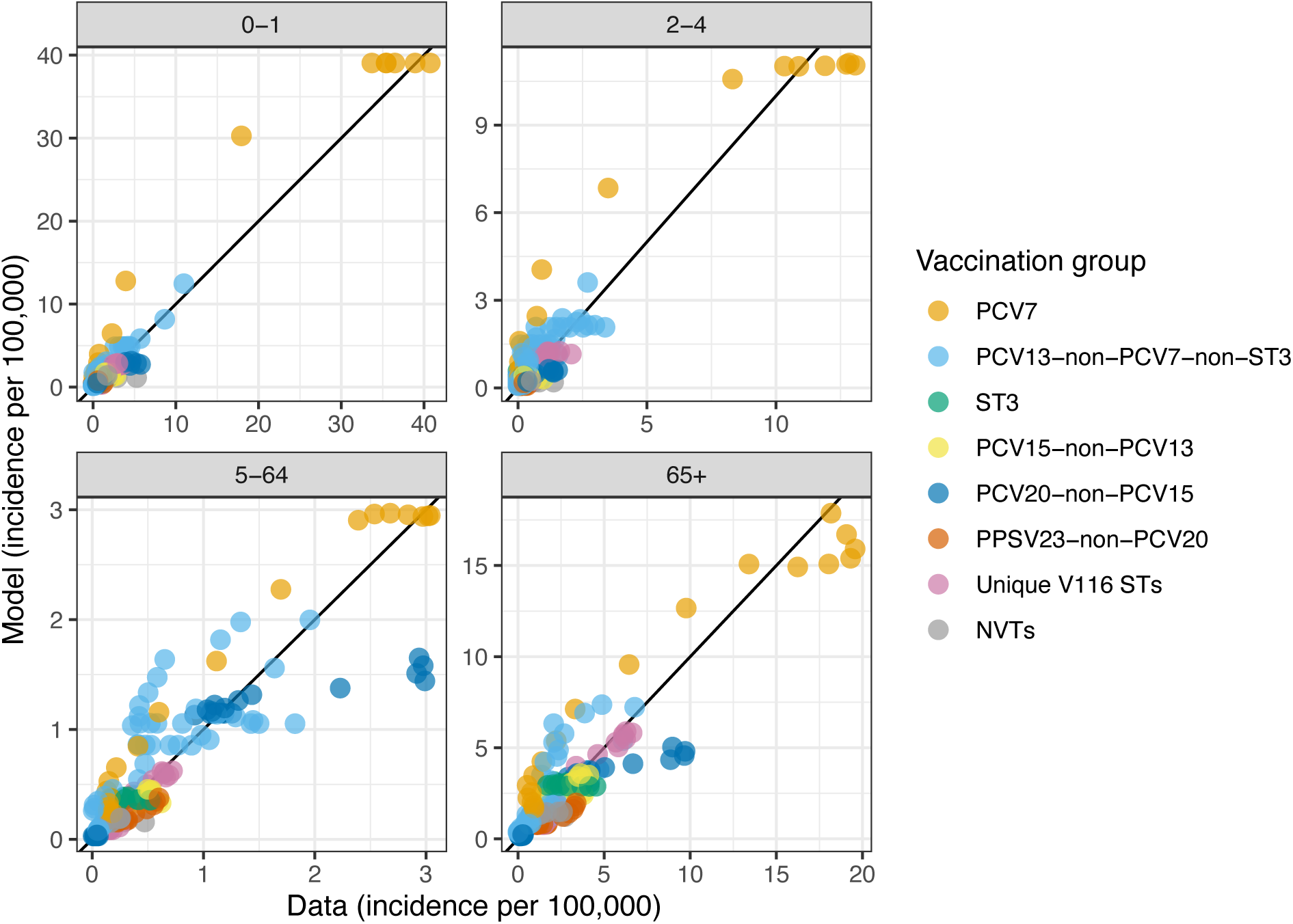
Model validation of calibrated DTM comparing data to model fit during the calibration period faceted by age group. Points denote annual and STC-specific incidence and are colored by vaccination grouping. Black line indicates a 1:1 line. The root mean square error (RMSE) are 1.47, 0.597, 0.312, and 1.29 for the 0-1, 2-4, 5-64, and 65+ age groups.

**Figure S5.**
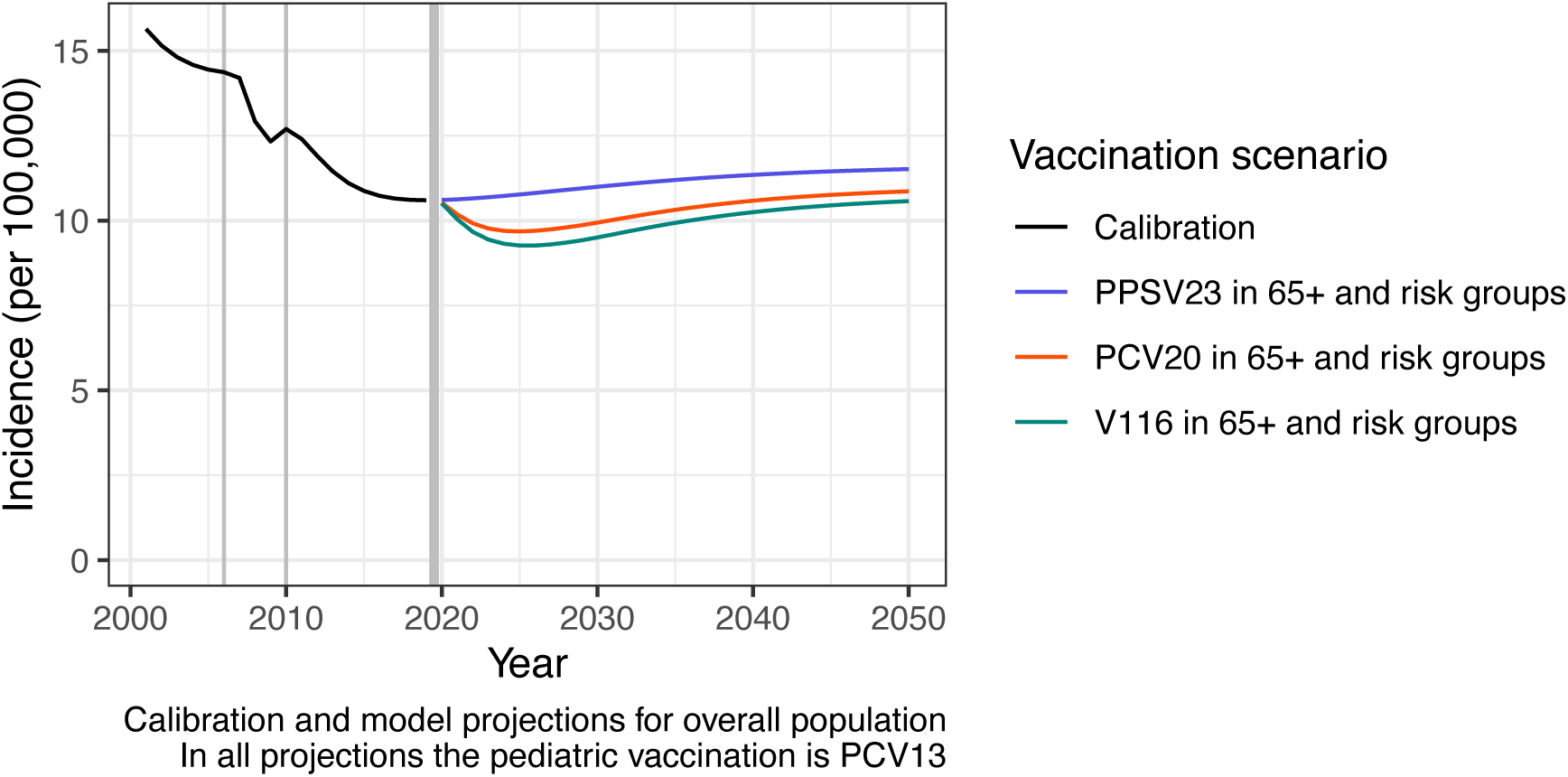
30-year projections of IPD incidence aggregated over serotype and age for three different adult vaccination scenarios. The black line denotes the calibrated model. The blue line denotes a scenario with PPSV23 in 65+ year-olds and risk groups, the red line denotes a scenario with PCV20 in 65+ year-olds and risk groups, and the teal line denotes a scenario with V116 in 65+ year-olds and risk groups. All projections assumed continued PCV13 vaccination in the pediatric population. Vertical grey lines indicate when different vaccines were introduced, PCV7 in 2006, PCV13 in 2010, and new vaccinations introduced in 2020.

**Figure S6.**
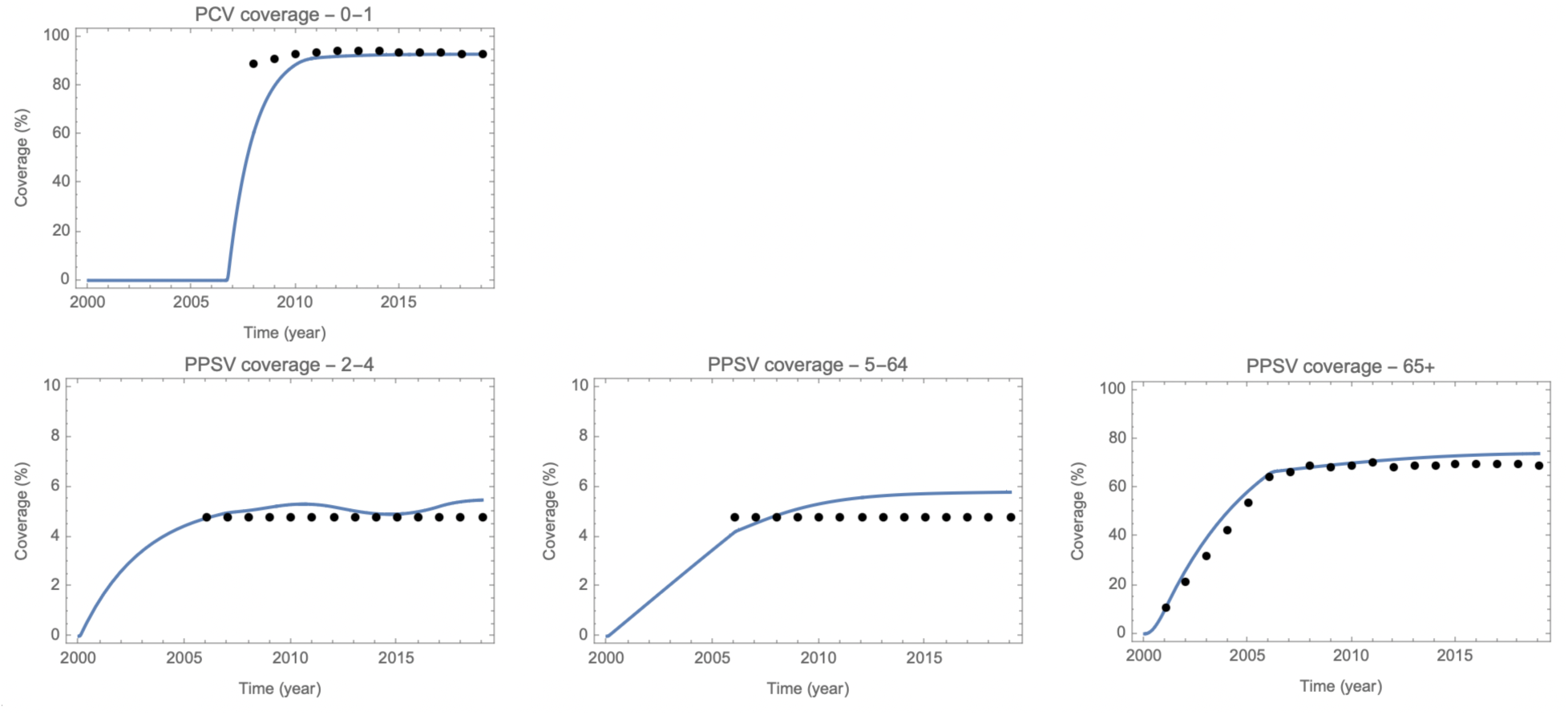
Calibrated vaccination coverage. Data (points) and model fits (solid line) are for 0-1-year-olds for PCVs and for 2-4-, 5-64-, and 65+ year-olds for PPSV.

**Figure S7.**
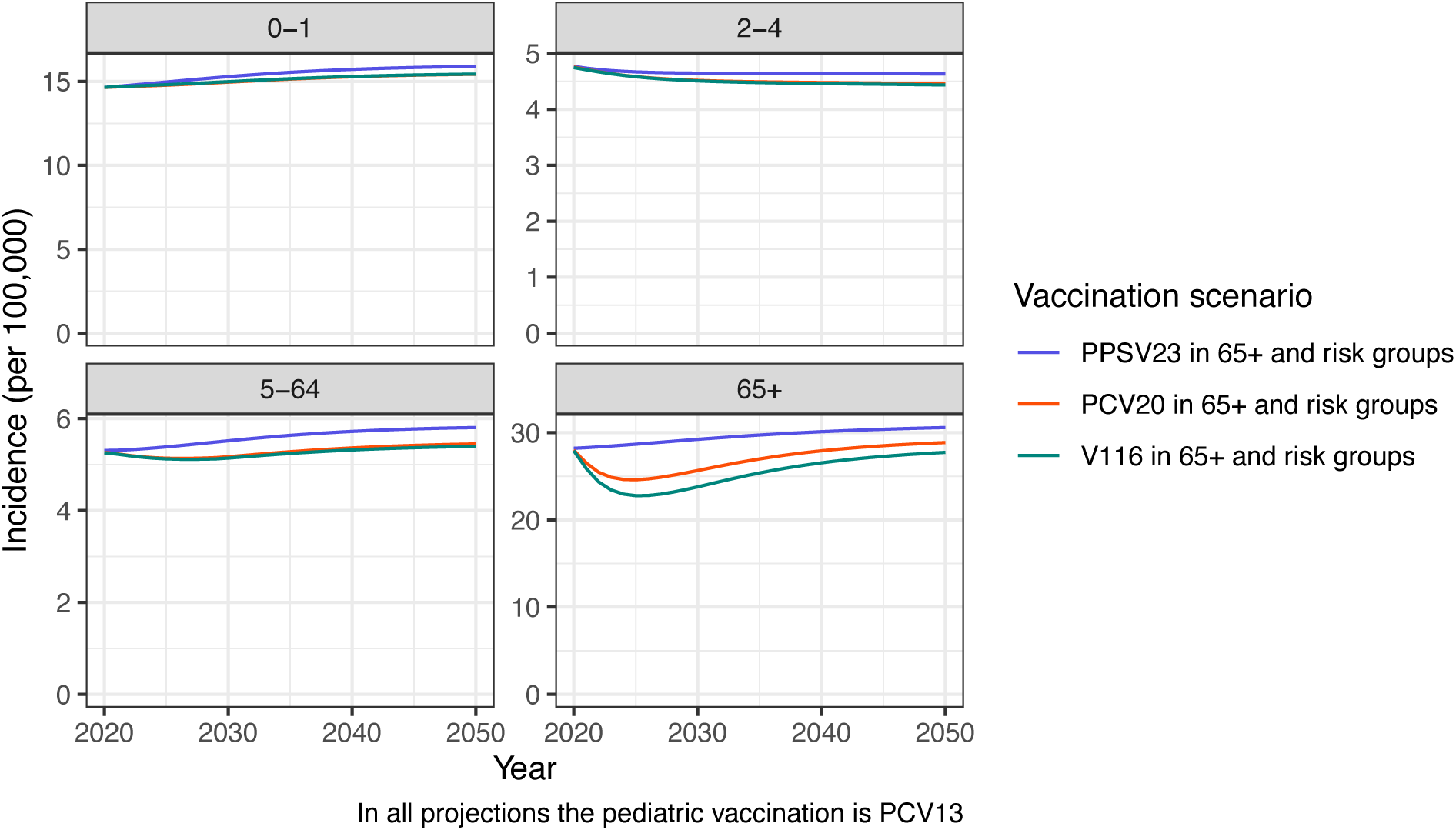
30-year projections of IPD incidence aggregated over serotypes for three different adult vaccination scenarios by age group. The blue line denotes a scenario with PPSV23 in 65+ year-olds and risk groups, the red line denotes a scenario with PCV20 in 65+ year-olds and risk groups, and the teal line denotes a scenario with V116 in 65+ year-olds and risk groups. All projections assumed continued PCV13 vaccination in the pediatric population.

**Figure S8.**
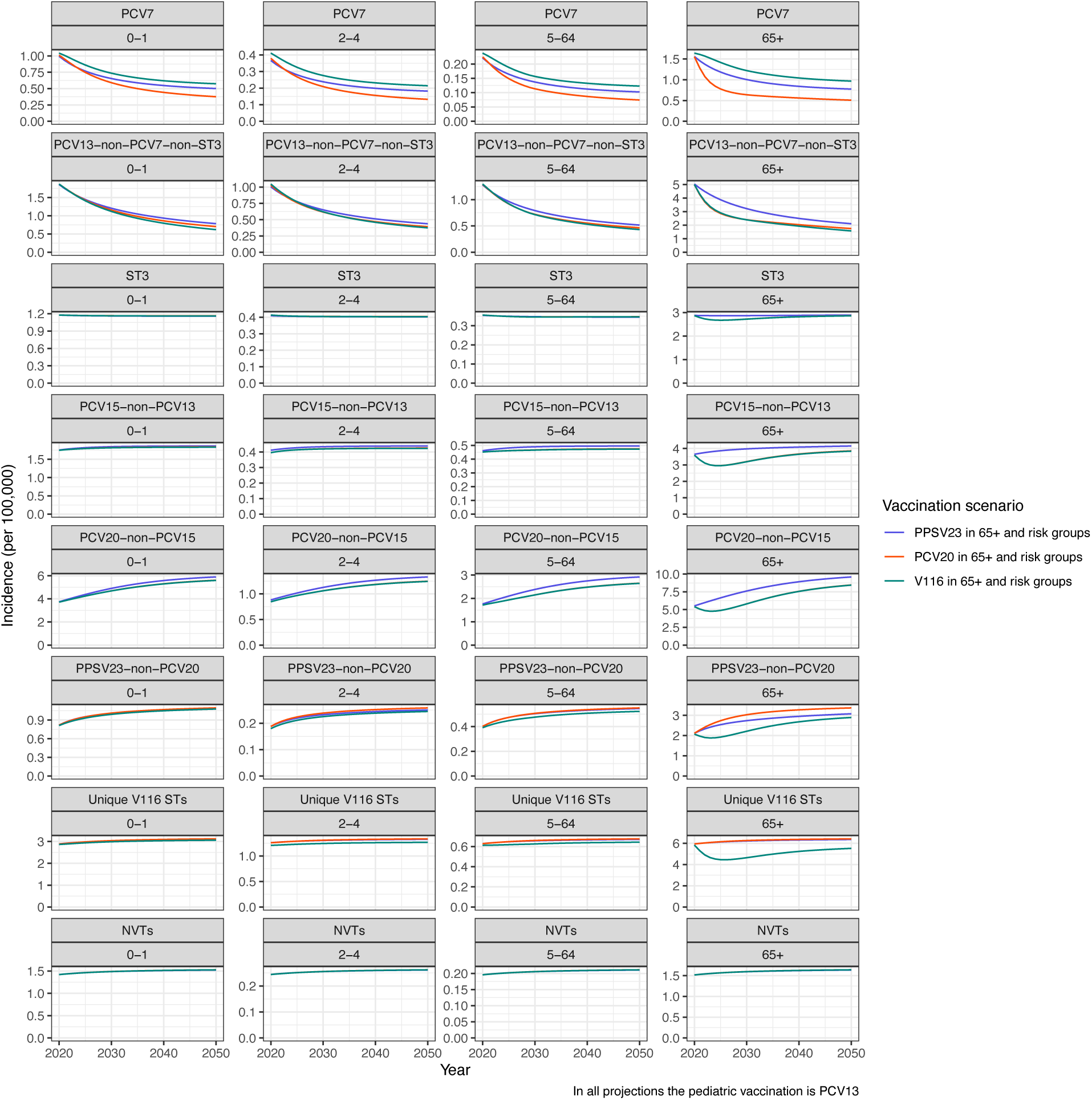
30-year projections of IPD incidence under three different adult vaccination scenarios by vaccination group and age group. The blue line denotes a scenario with PPSV23 in 65+ year-olds and risk groups, the red line denotes a scenario with PCV20 in 65+ year-olds and risk groups, and the teal line denotes a scenario with V116 in 65+ year-olds and risk groups. All projections assumed continued PCV13 vaccination in the pediatric population.

**Figure S9.**
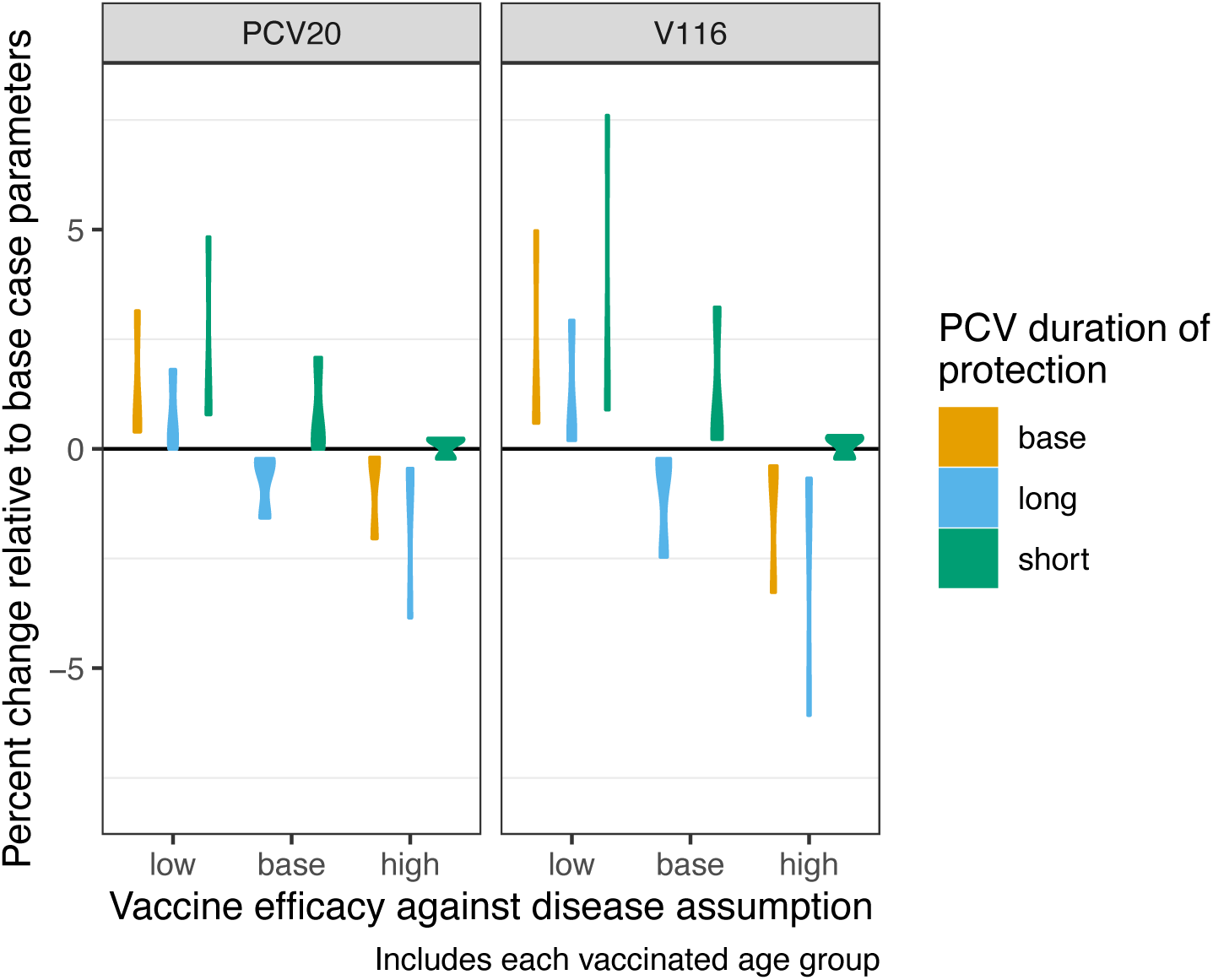
Distribution of percent change relative to base case parameters when varying the vaccine efficacy against disease and PCV duration of protection for each age group. Violin plots represent distribution over age. Results are faceted by PCV20 and V116 vaccination. Colors denote PCV duration of protection assumption. The base case parameters are the base duration of protection and base vaccine efficacy against disease (Table S7).

**Figure S10.**
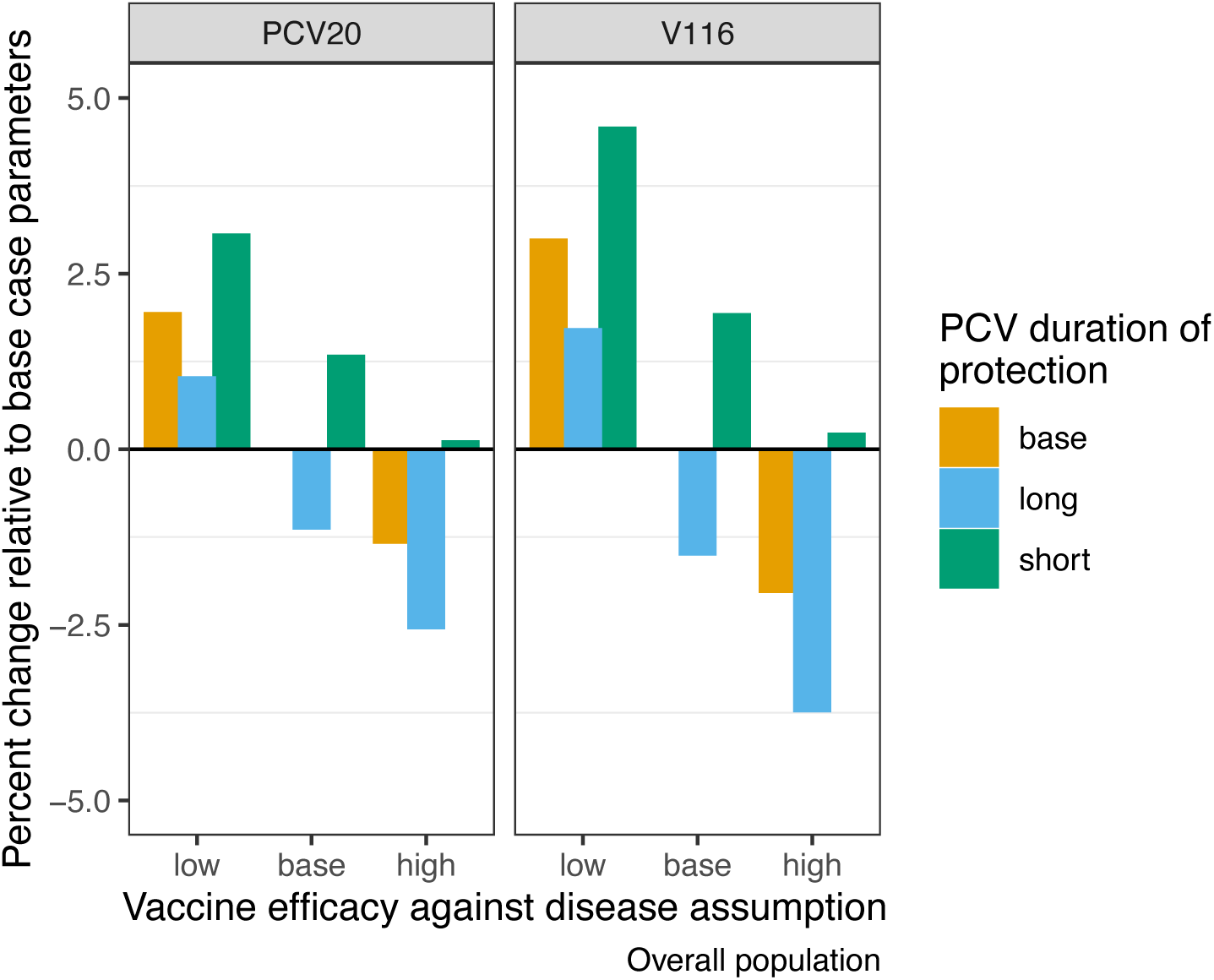
Percent change relative to base case parameters when varying the vaccine efficacy against disease and PCV duration of protection for the overall population. Results are faceted by PCV20 and V116 vaccination. Colors denote PCV duration of protection assumption. The base case parameters are the base duration of protection and base vaccine efficacy against disease (Table S7).

**Figure S11.**
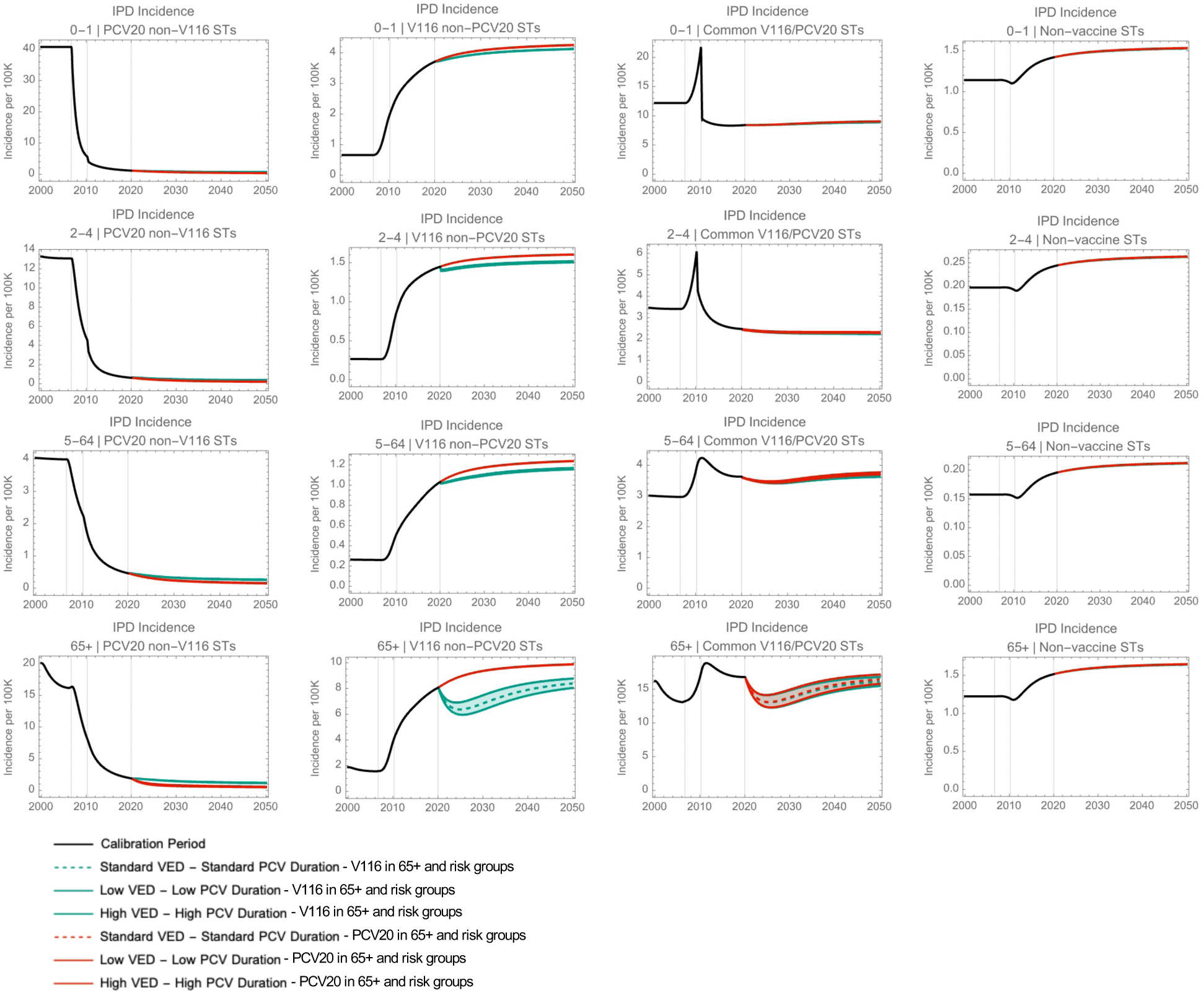
30-year projections from the sensitivity analysis by age group for each of the V116 groupings. Projections are grouped such that the base value of VE against disease is paired with the base duration of protection, the lower range of VE against disease is paired with the lower range for duration of protection, and the upper range of VE against disease is paired with the upper range of duration of protection. Vertical grey lines indicate when different vaccines were introduced, PCV7 in 2006, PCV13 in 2010, and new vaccinations introduced in 2020.

### Supplementary tables

**Table S4.** PCV Coverage for the 0-1-year-old age group [4].

| Year | VCR | Age group |
| --- | --- | --- |
| 2005 |  | 0-1 |
| 2006 |  | 0-1 |
| 2007 | 89.1 | 0-1 |
| 2008 | 90.1 | 0-1 |
| 2009 | 92.9 | 0-1 |
| 2010 | 93.6 | 0-1 |
| 2011 | 94.2 | 0-1 |
| 2012 | 94.4 | 0-1 |
| 2013 | 94.1 | 0-1 |
| 2014 | 93.9 | 0-1 |
| 2015 | 93.5 | 0-1 |
| 2016 | 93.5 | 0-1 |
| 2017 | 93.3 | 0-1 |
| 2018 | 92.8 | 0-1 |
| 2019 | 93.2 | 0-1 |

VCR = vaccination coverage rate

**Table S5.** PPSV23 Coverage for the 65+ year-old age group. Data were from 2005-2018. For 2019, we assumed the same coverage as 2018 [5].

| Year | VCR | Age groups |
| --- | --- | --- |
| 2005 | 64.4 | 65+ |
| 2006 | 66.6 | 65+ |
| 2007 | 69 | 65+ |
| 2008 | 68.2 | 65+ |
| 2009 | 69.4 | 65+ |
| 2010 | 70.5 | 65+ |
| 2011 | 68.3 | 65+ |
| 2012 | 69.1 | 65+ |
| 2013 | 68.9 | 65+ |
| 2014 | 69.8 | 65+ |
| 2015 | 70.1 | 65+ |
| 2016 | 69.8 | 65+ |
| 2017 | 69.5 | 65+ |
| 2018 | 69.2 | 65+ |
| 2019 | 69.2 | 65+ |

VCR = vaccination coverage rate

**Table S6.** PPSV23 Coverage for the 2-4- and 5-64-year-old age group. These calculations assume that the VCR of clinical risk groups is 49.0% and that the proportion of the 2-64-year-old population that falls within a clinical risk group is 9.7% [6].

| Year | VCR | Age groups |
| --- | --- | --- |
| 2005 | 4.78 | 2-4 and 5-64 |
| 2006 | 4.78 | 2-4 and 5-64 |
| 2007 | 4.78 | 2-4 and 5-64 |
| 2008 | 4.78 | 2-4 and 5-64 |
| 2009 | 4.78 | 2-4 and 5-64 |
| 2010 | 4.78 | 2-4 and 5-64 |
| 2011 | 4.78 | 2-4 and 5-64 |
| 2012 | 4.78 | 2-4 and 5-64 |
| 2013 | 4.78 | 2-4 and 5-64 |
| 2014 | 4.78 | 2-4 and 5-64 |
| 2015 | 4.78 | 2-4 and 5-64 |
| 2016 | 4.78 | 2-4 and 5-64 |
| 2017 | 4.78 | 2-4 and 5-64 |
| 2018 | 4.78 | 2-4 and 5-64 |
| 2019 | 4.78 | 2-4 and 5-64 |

VCR = vaccination coverage rate

**Table S7.** Vaccine efficacy against disease for the 0-1-year-old age group [7].

| Serotype class (STs) | VE against disease (PCV7) | VE against disease (PCV13) |
| --- | --- | --- |
| 1 (4, 6B, 9V, 14, 18C, 19F, 23F) | 0.96 | 0.96 |
| 2 (1, 5) | 0 | 0.84 |
| 3 (3) | 0 | 0.41 |
| 4 (7F, 19A) | 0 | 0.93 |
| 5 (6A, 6C) | 0 | 0.96 |
| 6 (22F, 33F) | 0 | 0 |
| 7 (9N, 17F, 20) | 0 | 0 |
| 8 (8, 10A, 11A, 12F) | 0 | 0 |
| 9 (15B) | 0 | 0 |
| 10 (15A, 15C, 16F, 23A, 23B, 24F, 31, 35B) | 0 | 0 |
| 11 (NVTs) | 0 | 0 |

VE = vaccine efficacy; NVTs = non-vaccine types

**Table S8.** Vaccine efficacy against disease for the 2-4-year-old and 5-64-year-old risk group and 65+ year-old population for PPSV23 [8], V116 [7], and PCV20 [7]. Note that given the lack of real-world evidence for vaccine effectiveness against disease for novel vaccine serotypes (STCs 6-10), we assumed equivalency with previous VTs in adult PCVs.

| Serotype class (STs) | VE against disease (PPSV23) | VE against disease (V116) | VE against disease (PCV20) |
| --- | --- | --- | --- |
| 1 (4, 6B, 9V, 14, 18C, 19F, 23F) | 0.45 | 0 | 0.75 |
| 2 (1, 5) | 0.23 | 0 | 0.75 |
| 3 (3) | 0.22 | 0.26 | 0.26 |
| 4 (7F, 19A) | 0.59 | 0.75 | 0.75 |
| 5 (6A, 6C) | 0 | 0.75 | 0.75 |
| 6 (22F, 33F) | 0.54 | 0.75 | 0.75 |
| 7 (9N, 17F, 20) | 0.65 | 0.75 | 0 |
| 8 (8, 10A, 11A, 12F) | 0.57 | 0.75 | 0.75 |
| 9 (15B) | 0.53 | 0 | 0.75 |
| 10 (15A, 15C, 16F, 23A, 23B, 24F, 31, 35B) | 0 | 0.75 | 0 |
| 11 (NVTs) | 0 | 0 | 0 |

VE = vaccine efficacy; NVTs = non-vaccine types

**Table S9.** Parameters varied in the DSA, including base case values with lower and upper ranges.

| Parameter description | Base value | Lower range | Upper range |
| --- | --- | --- | --- |
| IPD VE STC1 | 0.75 | 0.6 | 0.85 |
| IPD VE STC2 | 0.75 | 0.6 | 0.85 |
| IPD VE STC3 | 0.26 | 0.21 | 0.29 |
| IPD VE STC4 | 0.75 | 0.6 | 0.85 |
| IPD VE STC5 | 0.75 | 0.6 | 0.85 |
| IPD VE STC6 | 0.75 | 0.6 | 0.85 |
| IPD VE STC7 | 0.75 | 0.6 | 0.85 |
| IPD VE STC8 | 0.75 | 0.6 | 0.85 |
| IPD VE STC9 | 0.75 | 0.6 | 0.85 |
| IPD VE STC10 | 0.75 | 0.6 | 0.85 |
| IPD VE STC11 | 0.75 | 0.6 | 0.85 |
| PCV duration of protection in adults | 10 years | 8 years | 12 years |

IPD = invasive pneumococcal pneumonia; VE = vaccine efficacy; STC = serotype class

**Table S10.**
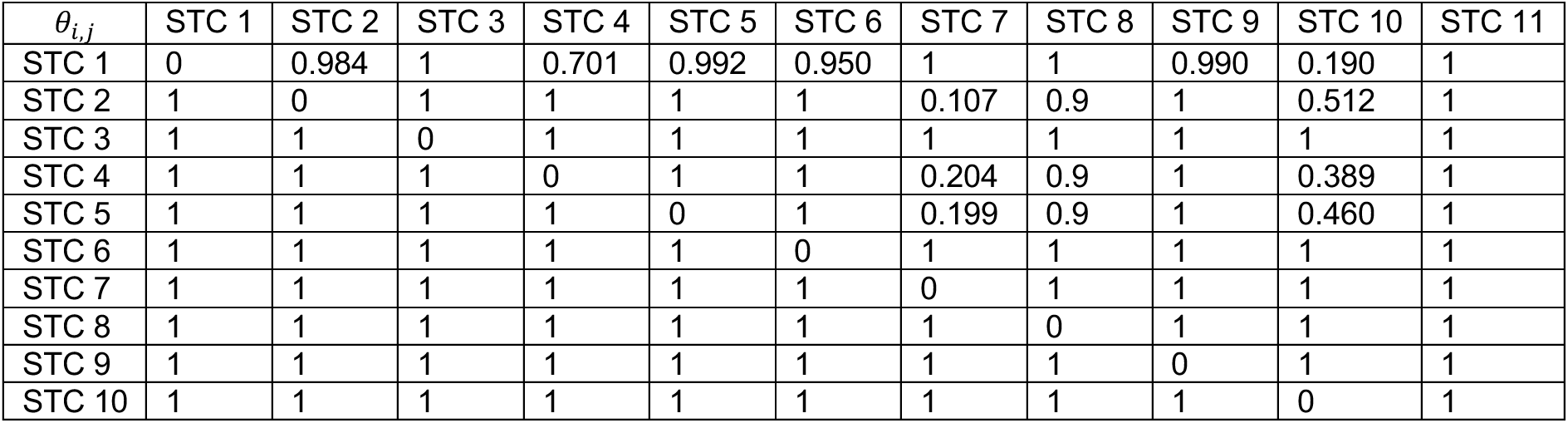
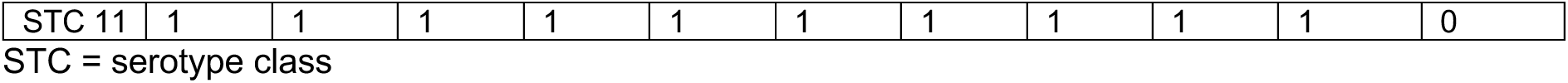
Calibrated parameters for serotype competition, *θ*_*i*,*j*_. The likelihood of acquisition of a second STC (*j*, represented by columns) if currently colonized with another STC (*i*, represented by rows).

**Table S11.**
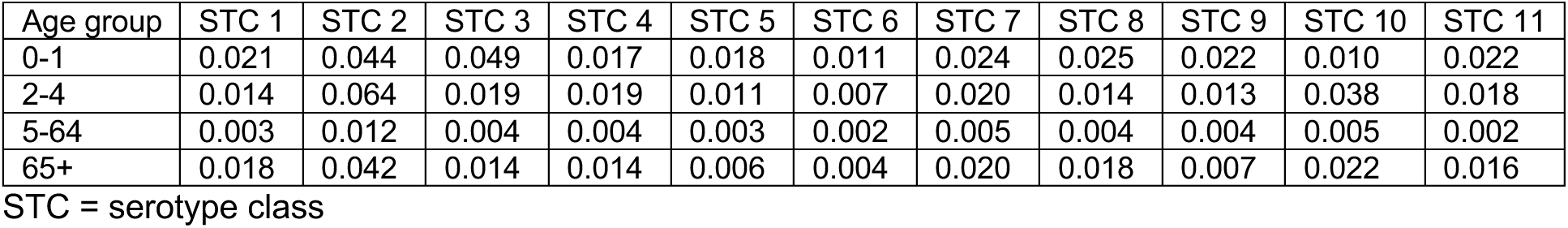
Calibrated parameters for probability of carriage acquisition, *β*_*a*,*i*_. Carriage acquisition for STC *i* (represented by columns) per contact in age group *a* (represented by rows).

**Table S12.**
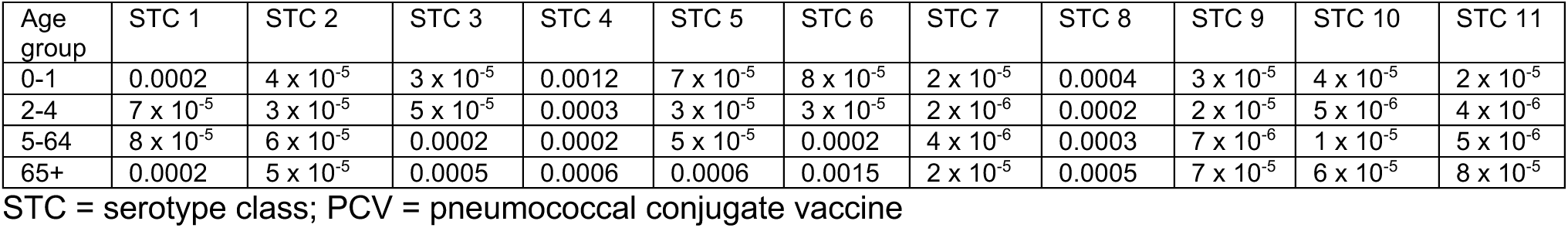
Calibrated parameters for probability of disease given colonization, i.e., the case-to-carrier ratio, *ρ*_*a*,*i*_. Probability of disease by STC *i* (represented by columns) in age group *a* (represented by rows).

**Table S13.**
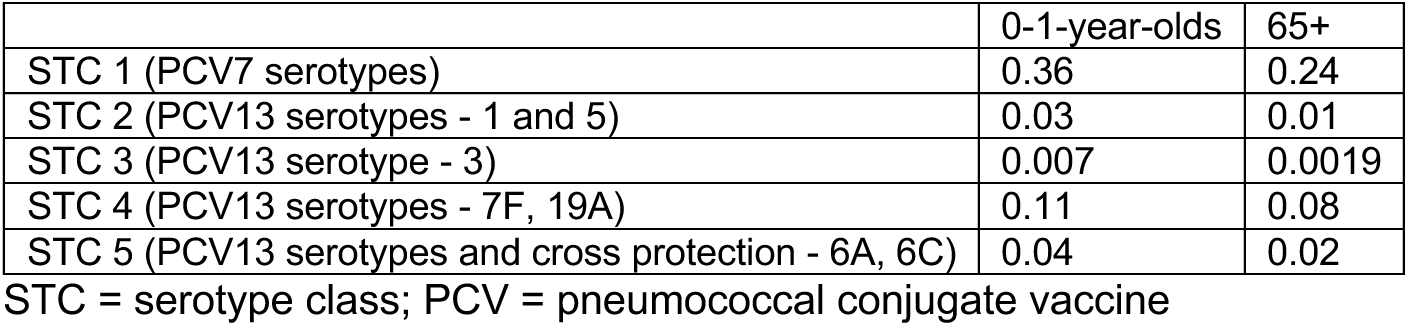
Calibrated parameters for vaccine efficacy against carriage acquisition, *∈*_*σ*,*a*,*i*_.

|  | 0-1-year-olds | 65+ |
| --- | --- | --- |
| STC 1 (PCV7 serotypes) | 0.36 | 0.24 |
| STC 2 (PCV13 serotypes - 1 and 5) | 0.03 | 0.01 |
| STC 3 (PCV13 serotype - 3) | 0.007 | 0.0019 |
| STC 4 (PCV13 serotypes - 7F, 19A) | 0.11 | 0.08 |
| STC 5 (PCV13 serotypes and cross protection - 6A, 6C) | 0.04 | 0.02 |

STC = serotype class; PCV = pneumococcal conjugate vaccine

